# Network responses to meditation and baseline connectivity correlates of cognitive improvement in schizophrenia

**DOI:** 10.1101/2025.11.20.25340575

**Authors:** Qing Wang, Lizhao Du, Jialing Sheng, Qiao Wang, Yuan Shi, Ting Xue, Zhong Sun, Yingying Tang, Donghong Cui

**Affiliations:** Shanghai Key Laboratory of Psychotic Disorders, Brain Health Institute, National Center for Mental Disorders, Shanghai Mental Health Center, School of Medicine, Shanghai Jiao Tong University, Shanghai, 200030, China; NeuroImaging Core, Shanghai Mental Health Center, School of Medicine, Shanghai Jiao Tong University, 600 S. Wanping Rd., Shanghai, 200030, China; Shanghai Med-X Engineering Research Center, School of Biomedical Engineering, Shanghai Jiao Tong University, Shanghai, China; The First Minzheng Mental Health Center, No. 9999 Zhongchun Rd., Minhang, Shanghai, 201105, China; Department of Public Health, Brody School of Medicine, East Carolina University, 610 Moye Blvd, Mailstop 660, Greenville, NC 27834, U.S.A; Brain Science and Technology Research Center, Shanghai Jiao Tong University, Shanghai, China

## Abstract

Meditation-based rehabilitation may improve symptoms and cognition in schizophrenia, but how these responses relate to changes in brain-network interactions remains unclear. We conducted a secondary analysis of longitudinal clinical assessments and resting-state functional MRI at baseline, 3 and 8 months in a randomized trial of 64 male inpatients comparing meditation-based rehabilitation with general rehabilitation. Each analysis included all participants with the required measurements: 62 for clinical, 44 for three-visit connectivity and 43 for joint neural–clinical models. Symptom treatment effects increased between months 3 and 8, whereas cognitive benefit was clearest at 3 months. In the clinical cross-lagged panel model, early cognitive improvement was associated with cumulative symptom improvement at 8 months (β = 0.136; nominal P = 0.044); bootstrap intervals included zero for this association and the indirect treatment effect. Intrinsic network properties showed no treatment contrasts surviving multiplicity correction. Spectral dynamic causal modelling and parametric empirical Bayes identified eight treatment-associated connections shared across models with and without head-motion adjustment. Two early effects were consistent across five model specifications, sharing a supramarginal–frontal source with sensorimotor and bilateral middle-frontal targets. Adding baseline connectivity and early changes in these connections to clinical and motion covariates increased conditional in-sample R² most for 3-month cognitive improvement, by 0.147. Baseline connectivity supplied most of this increment (0.125); its two associations, modelled as common across treatment arms, retained their directions across all participant omissions. Early-change associations with clinical outcomes were imprecise. These findings distinguish treatment-responsive network interactions from the baseline connectivity that adds most information about cognitive improvement. They motivate a testable account in which initial sensory–attention coordination supports engagement with rehabilitation, while changing practice demands shape network responses.

## Introduction

Meditation-based interventions offer a way to train attention and responses to internal experience alongside established mental health care. Their clinical relevance is supported by evidence across several distinct settings: mindfulness-based cognitive therapy reduces relapse risk in recurrent depression, and mindfulness-based stress reduction was noninferior to escitalopram for anxiety severity in a randomized trial. [1, 2] Broader syntheses identify benefits for mental wellbeing and psychological distress, including outside clinical services. [3, 4] The size of these benefits depends on the condition, programme and comparator, with more modest effects against active interventions. [5–7] Establishing how meditation affects clinically relevant processes is therefore important for understanding where, when and for whom it may contribute to care.

Schizophrenia presents a particularly consequential setting for this question because persistent psychotic experiences and cognitive difficulties can coexist during ongoing treatment. Adapted mindfulness-informed programmes have improved selected symptom, functional and hospitalization outcomes, although their effects vary across endpoints and they commonly combine meditation with psychoeducation or cognitive therapy. [8–10] In our parent randomized trial, an intensive meditation-based rehabilitation programme improved persistent hallucinations and delusions and quality of life in male inpatients with chronic schizophrenia. [11] Cognitive improvement is also a plausible treatment target: quantitative syntheses support benefits in selected cognitive domains, with a newer, larger review finding effects on several accuracy-based measures against active controls. [12, 13] How cognitive and symptom responses evolve together, and what changes in brain function contribute to understanding those responses, remain clinically meaningful questions.

Mechanistic accounts of meditation emphasize attentional regulation, bodily awareness and the regulation of emotional and self-related processing. [14, 15] These accounts implicate coordination among distributed functional systems. Resting-state independent component analysis can identify spatially distributed networks, providing a data-driven unit for studying their interactions. [16] Task-based fMRI has separated mind wandering, awareness, reorienting and sustained focus during meditation, identified altered cortical responses during interoceptive attention, and linked mindfulness training to attention-related responses during emotion regulation. [17–19] Functional imaging meta-analysis also indicates that different practices recruit partly distinct patterns. [20] Such findings provide candidate operations through which training could act. A central challenge is to connect these operations to changes in ongoing brain organization, their persistence, and the outcomes that matter to patients.

Resting-state studies complement task experiments by examining ongoing interactions among networks, although acquisition instructions range from unconstrained rest to deliberate sensory attention. Findings involving default-mode, control, attention and sensory systems have been reported in experienced meditators and following training. [21, 22] Randomized work has linked altered resting connectivity to subsequent inflammatory outcomes and described changes in executive-control network coupling. [23, 24] A meditation-versus-relaxation trial has also used spectral dynamic causal modelling (DCM) to characterize changes in directed pons–posterior cingulate coupling. [25] These studies establish important precedents, while differences in population, mental state, intervention and follow-up constrain transfer to chronic psychiatric illness. Longitudinal observations are especially informative: in one mindfulness-based stress reduction trial, a posterior cingulate–prefrontal connectivity effect observed after training was not maintained at later follow-up. [26] Repeated assessments can therefore distinguish an early-peaking response from a sustained shift in network coupling, a distinction that a single post-treatment scan would leave unresolved.

The remaining question extends beyond detecting a treatment-related brain difference. Initial neural organization and subsequent neural change can make different contributions to clinical improvement. Baseline coupling may capture characteristics relevant to engagement with rehabilitation, whereas treatment-associated changes may reflect adaptation during practice. An intervention study can examine both quantities and assess their correspondence with cognitive and symptom outcomes. This approach addresses the call for longitudinal, actively controlled and theoretically explicit mindfulness research. [27] It also allows more specific cognitive hypotheses, including the distinction between increased processing capacity and greater efficiency through reduced interference. [28]

Previous publications from this trial established clinical efficacy and examined longitudinal insula-seed functional connectivity, regional gray-matter trajectories and white-matter structural networks. [11, 29–31] The insula study provided a healthy-control reference and followed meditation-treated patients; the gray-matter and diffusion-MRI studies retained the randomized rehabilitation comparison. The present analysis addresses a complementary functional systems question: how directed interactions among distributed networks evolve relative to the randomized comparator, and how baseline connectivity and early connectivity changes contribute to a joint account of cognitive and symptom improvement. We analysed clinical assessments and resting-state fMRI at baseline and at 3 and 8 months, combining time-specific treatment estimates, clinical cross-lagged panel models (CLPMs), intrinsic network measures, hierarchical DCM–parametric empirical Bayes analysis, and focused neural–clinical CLPMs. This secondary analysis aims to identify treatment-responsive circuitry and define the clinical information it provides, generating specific hypotheses for subsequent mechanistic tests.

## Results

### Clinical improvement and its temporal relationships

The trial randomized 64 male inpatients with chronic schizophrenia to a meditation-based programme or general rehabilitation (32 participants per group). The present complete-case clinical analysis included 62 participants (32 meditation and 30 control), assessed at baseline and at 3 and 8 months (Fig. 1a). This report extends the parent trial by examining time-specific average treatment effects, clinical–cognitive relationships and longitudinal network responses. [11] Analysis samples were determined by availability of the measurements required for each model: the complete longitudinal DCM sample comprised 44 participants, and the joint neural–clinical models included 43 participants with the required clinical data (Supplementary Table 1).

**Fig. 1.**
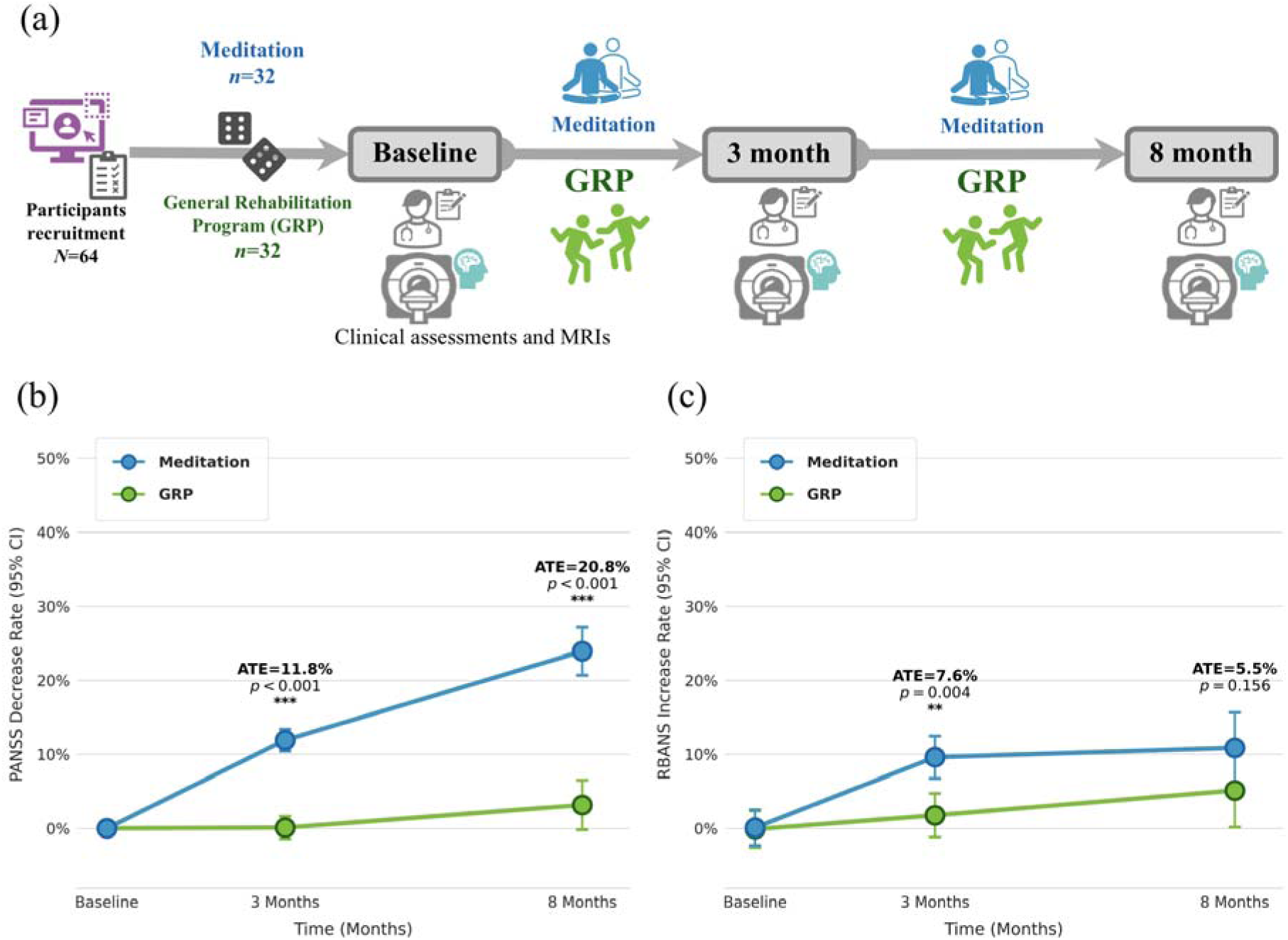
Trial design and longitudinal symptom and cognitive responses. **a,** Sixty-four male participants with chronic schizophrenia were randomized to a meditation-based programme or general rehabilitation programme (GRP; 32 per group). Clinical assessments and MRI were scheduled at baseline and after 3 and 8 months during the trial. **b,c,** Adjusted trajectories of PANSS decrease rate (b) and RBANS increase rate (c) in the complete-case clinical sample (n = 62; 32 meditation, 30 GRP; 186 observations). PANSS decrease rate was 100 × (baseline − follow-up)/(baseline − 30); RBANS increase rate was 100 × (follow-up − baseline)/baseline. Points are estimated marginal means at the pooled baseline-score mean; bars are model-based 95% confidence intervals. Mixed-effects models included treatment, categorical visit, their interaction and the corresponding baseline score, with participant-specific elapsed-time random slopes and no random intercept, fitted by restricted maximum likelihood. Average treatment effects (ATEs) are model-estimated meditation-minus-GRP group-by-visit contrasts relative to baseline. Annotations use percentage notation for absolute contrasts multiplied by 100; the values are interpreted in percentage points. Tests are two-sided model-based Wald tests without multiplicity correction; **P < 0.01, ***P < 0.001. Blue denotes meditation and green denotes GRP. PANSS, Positive and Negative Syndrome Scale; RBANS, Repeatable Battery for the Assessment of Neuropsychological Status.

Meditation increased PANSS decrease rate by 11.81 percentage points at 3 months (95% confidence interval (CI), 9.32–14.30; P < 0.001) and 20.77 percentage points at 8 months (15.94–25.60; P < 0.001; Fig. 1b). The corresponding average treatment effects on RBANS increase rate were 7.62 percentage points at 3 months (2.40–12.84; P = 0.004) and 5.52 percentage points at 8 months (−2.10–13.13; P = 0.156; Fig. 1c). Thus, symptom benefit was evident at both visits, whereas the later cognitive estimate remained compatible with negligible as well as positive effects. Follow-up-only models with participant-clustered uncertainty supported an increase in the symptom treatment contrast between months 3 and 8 but did not establish a decline in the cognitive contrast (Supplementary Tables 2 and 3).

In the clinical cross-lagged panel model (CLPM), both symptom and cognitive improvement showed positive autoregressive associations between months 3 and 8 (standardized coefficients β = 0.546 and 0.563, respectively; both P < 0.001; Fig. 2). Early cognitive improvement was associated with cumulative 8-month symptom improvement after adjustment for early symptoms, treatment and baseline covariates (β = 0.136; nominal P = 0.044). The reverse cross-lagged association was imprecise (β = 0.039; P = 0.829). The cognitive-to-later-symptom association was also imprecise under the corresponding participant-bootstrap regression interval (95% CI, −0.050–0.305). Exploratory mediation analysis yielded a standardized indirect-effect estimate of 0.043 for treatment → early cognition → later symptom improvement (95% participant-bootstrap CI, −0.015–0.150). The observations therefore motivate investigation of a clinical–cognitive relationship (Supplementary Tables 4 and 5).

**Fig. 2.**
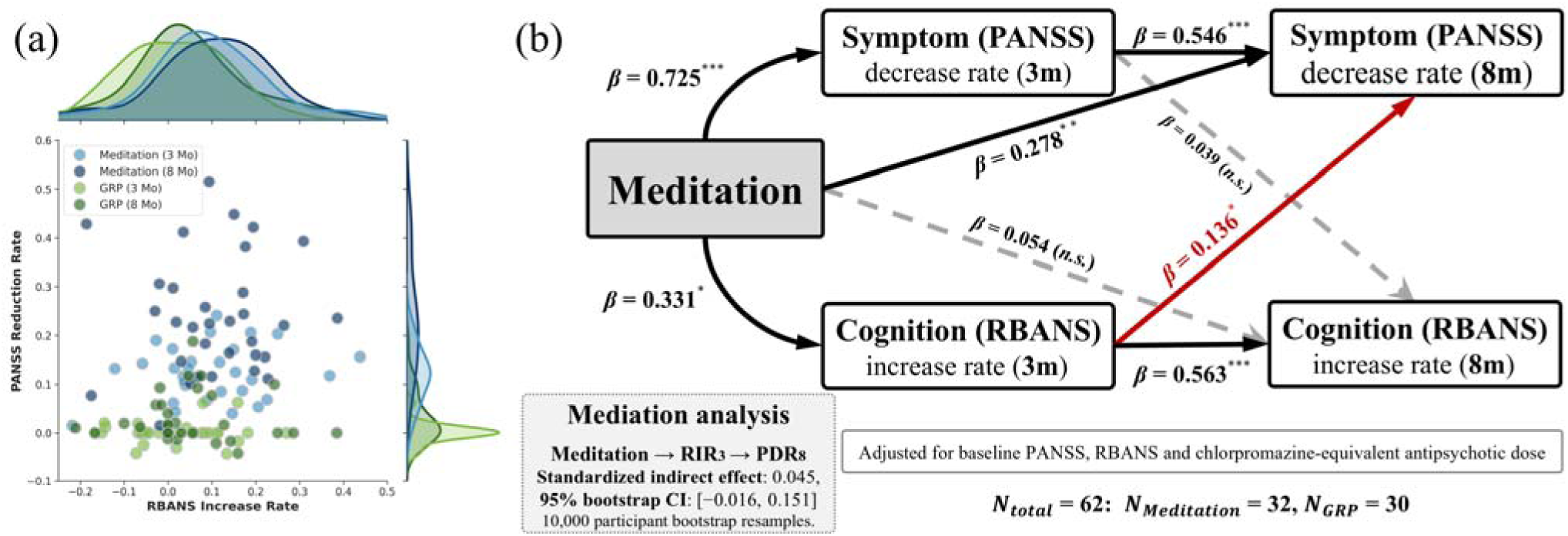
Temporal clinical–cognitive associations and exploratory mediation. **a,** Joint distributions of cumulative RBANS increase rate and PANSS decrease rate after months 3 and 8. Points are participant–visit observations and marginal curves are kernel-density estimates. Blue and green identify meditation and GRP; lighter and darker shades identify months 3 and 8. Paired observations were available for 32 meditation and 30 GRP participants at each visit; PANSS marginal densities additionally include one GRP participant with available PANSS data. **b,** Cross-lagged panel model in 62 participants (32 meditation, 30 GRP). Arrows show standardized coefficients (β), adjusted for baseline PANSS, RBANS and chlorpromazine-equivalent dose. Treatment was also standardized. Within-visit residual covariances were estimated but omitted from the diagram. Solid/dashed arrows indicate nominal P < 0.05/P ≥ 0.05; *P < 0.05, **P < 0.01, ***P < 0.001, from two-sided native Wald tests. The red arrow highlights early cognition versus later cumulative symptom improvement, conditional on early symptoms and the other predictors. The mediation inset reports the standardized indirect-effect estimate from corresponding regression equations (a × b = 0.043; 95% percentile CI, −0.015–0.150), using 10,000 participant bootstrap resamples with original scaling fixed. The interval does not provide clear evidence of mediation. PDR, PANSS decrease rate; RIR, RBANS increase rate.

### Treatment-related changes in directed network interactions

We characterized both intrinsic network properties and directed interactions among nine independent components identified by canonical independent component analysis (ICA; Fig. 3a). Component labels combined dominant anatomical features with atlas-supported network affiliations, retaining the IC identifier to distinguish the empirical components from canonical networks. Intrinsic spatial expression, sample entropy, permutation entropy and the detrended fluctuation analysis exponent showed no baseline-adjusted between-arm contrast at months 3 or 8 surviving false-discovery-rate correction (minimum adjusted P = 0.1504; Fig. 3b; Supplementary Table 6). These results provide no clear evidence for treatment effects on the measured local properties; and they do not establish equivalence between treatment arms as well.

**Fig. 3.**
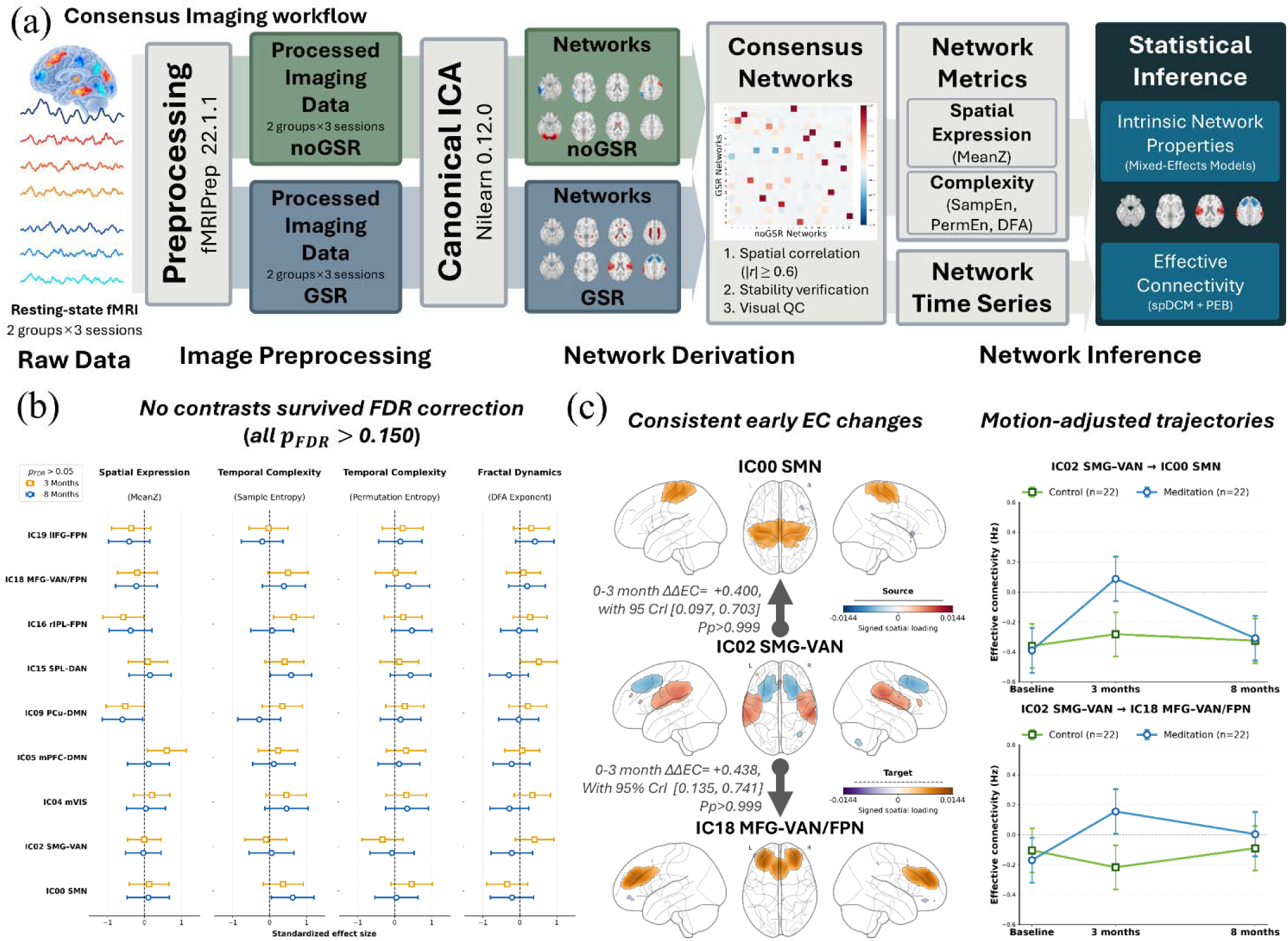
Imaging workflow, intrinsic properties and focal connectivity changes. **a,** Network analysis workflow. Components were retained using spatial correspondence across decompositions with and without global signal regression (|r| ≥0.60) and visual quality control. Intrinsic properties and directed effective connectivity (EC) were examined using mixed-effects models and spectral DCM with hierarchical PEB, respectively. **b,** Meditation-minus-control contrasts for spatial expression (MeanZ), sample entropy, normalized permutation entropy and DFA exponent at months 3 (gold squares) and 8 (blue circles). Points are coefficients in follow-up outcome-SD units; bars are nominal 95% Wald CIs. Models included participant intercepts and baseline measure, age and visit-specific FD. Benjamini–Hochberg correction was applied across nine components within each measure and visit; no contrast survived (minimum adjusted P = 0.1504). The local-measure sample preparation is specified in Supplementary Methods and Table 1. **c,** Two connections with positive early treatment-change contrasts whose 95% CrIs excluded zero across five specifications. Signed spatial maps use separate source/target scales. Arrow labels show meditation-minus-control baseline-to-month-3 change contrasts in Hz and full-PEB 95% CrIs. Pp refers separately to BMA support for treatment-by-quadratic-time terms. Trajectory points and bars are full-PEB fitted means and pointwise 95% CrIs evaluated at mean FD at all three visits (n = 44, 22 per arm; 132 scans). Lines connect visits. Focal selection followed the within-study sensitivity comparisons. DCM, dynamic causal modelling; PEB, parametric empirical Bayes; FD, framewise displacement.

Spectral dynamic causal modelling (spDCM) and hierarchical parametric empirical Bayes (PEB) identified eight directed connections shared between models with and without mean framewise displacement adjustment. All eight shared connections had posterior inclusion probabilities greater than 0.999 for a treatment-by-time term, exceeding the applied selection threshold of 0.95. Five were associated with the quadratic temporal term and three with the linear term. The eight connections linked supramarginal, sensorimotor, visual, parietal, default-mode and frontal components (Fig. 4; Supplementary Tables 7 and 8). The temporal terms describe components of a jointly fitted trajectory.

**Fig. 4.**
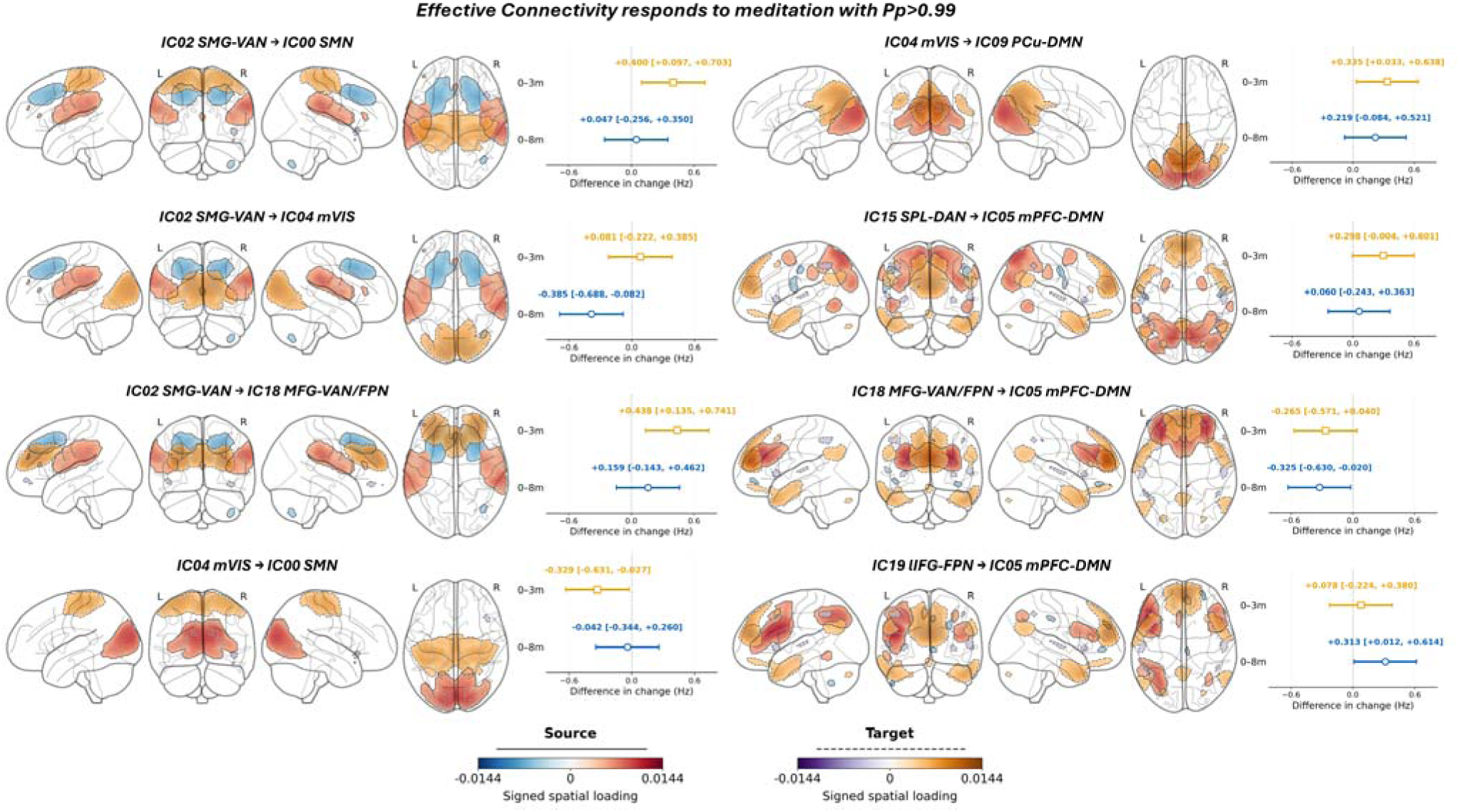
Eight treatment-associated connections shared across PEB specifications. Anatomical maps and treatment-change estimates for the eight directed connections selected in both reference and motion-adjusted PEB models. Selection required Bayesian model-averaging posterior probability Pp >0.95 for at least one treatment-by-linear or treatment-by-quadratic term; selected terms for all eight shared connections exceeded 0.999 in both specifications. Titles identify source and target components. Glass brains overlay signed component loadings with separate source/target colour scales and solid/dashed support contours. Display threshold: 20th percentile of absolute nonzero loadings (upper 80%). Gold squares and blue circles show meditation-minus-control differences in baseline-to-month-3 and baseline-to-month-8 EC change, respectively. Bars are pointwise 95% CrIs from the full PEB model adjusted for FD at all three visits (n = 44; 22 per arm; 132 scans). Labels give means and interval limits in Hz. Positive values indicate more positive EC change in meditation. Visit contrasts combine the linear and quadratic coefficients and their covariance; they are distinct from the BMA inclusion probabilities. Six FD-only candidates and complete sensitivity comparisons are provided in Supplementary Table 8. Component naming evidence and abbreviations appear in Supplementary Table 7.

Two early contrasts were particularly consistent across the specified model sensitivities: IC02 SMG–VAN → IC00 SMN and IC02 SMG–VAN → IC18 MFG–VAN/FPN. In the motion-adjusted model, the meditation-minus-control differences in baseline-to-month-3 change were 0.400 Hz (95% credible interval (CrI), 0.097– 0.703) and 0.438 Hz (0.135–0.741), respectively (Fig. 3c). Both remained positive with intervals excluding zero across the five fitted specifications. Their cumulative month-8 contrasts were less precise: 0.047 Hz (−0.256–0.350) and 0.159 Hz (−0.143–0.462). The motion-adjusted fitted trajectories accordingly showed their clearest separation at month 3. These sensitivity results motivated the focused neural–clinical models; the choice was based on treatment-related connectivity evidence.

The complete eight-connection display preserves effects of both signs and both follow-up contrasts (Fig. 4). Posterior support for inclusion of a temporal coefficient is distinct from the uncertainty in a particular baseline-to-visit contrast. Accordingly, not every 3-or 8-month interval excludes zero. We describe the two focal effects as consistent across the examined specifications.

### Baseline connectivity contributed most to the description of early cognitive improvement

We incorporated the two focal connections into the clinical model in 43 participants with complete longitudinal imaging, clinical and motion data (22 meditation and 21 control). Four joint models used identical participants and estimation conventions: the clinical reference model (M0), addition of visit-specific motion (M1), addition of both baseline ECs (M2), and addition of both early EC changes (M3; Fig. 5a). This sequence distinguishes the information associated with initial network organization from that associated with subsequent connectivity change.

**Fig. 5.**
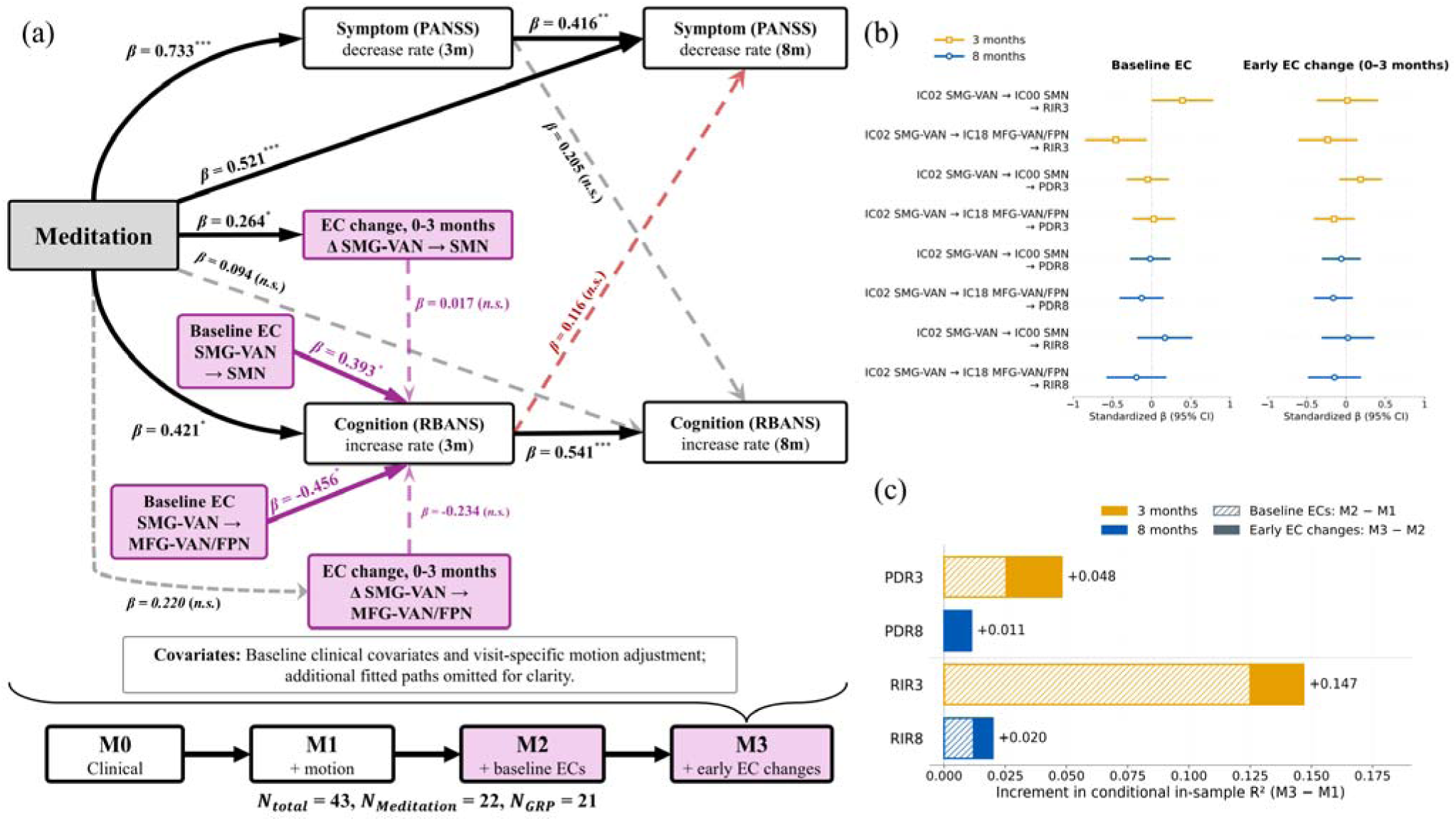
Neural–clinical associations and additional in-sample explained variance. All four neural–clinical CLPMs used the same 43 participants with complete required inputs (22 meditation, 21 control), with PDR and RIR measured as cumulative improvement from baseline. **a,** Focused representation of M3, showing clinical paths, treatment-to-EC-change paths and neural associations with early cognition. Neural inputs comprise baseline EC and month-3 minus baseline EC for IC02 SMG–VAN → IC00 SMN and IC02 SMG–VAN → IC18 MFG–VAN/FPN. Labels are standardized β. Solid/dashed paths indicate nominal P <0.05/P ≥0.05; *P <0.05, **P <0.01, ***P <0.001 (two-sided native Wald tests). Purple identifies neural variables; red highlights the clinical cognition-to-later-symptoms path. Covariates include baseline clinical scores and CPZ, FD0/FD3 in early equations and FD0/FD3/FD8 in late equations. Other fitted paths and residual covariances are omitted. M0 is the clinical reference; M1 adds motion, M2 baseline ECs and M3 early changes. **b,** All 16 jointly estimated neural–clinical coefficients. Bars are pointwise 95% native expected-information Wald CIs, rescaled with fitted SDs held fixed. Gold squares indicate month 3 and blue circles month 8. Intervals and P values are unadjusted for multiple comparisons or preceding EC selection. **c,** Increment in conditional in-sample R²: hatched baseline segments show M2 − M1, filled early-change segments M3 − M2, and labels the combined M3 − M1 increment. Gold/blue identify month 3/month 8. The ordered decomposition has no estimated ΔR² interval. It describes added in-sample information. PDR, PANSS decrease rate; RIR, RBANS increase rate; EC, effective connectivity.

Relative to the clinical-plus-motion model, the full neural model increased conditional in-sample R² by 0.048 for month-3 symptom improvement, 0.011 for month-8 symptom improvement, 0.147 for month-3 cognitive improvement and 0.020 for month-8 cognitive improvement (Fig. 5c). The largest increment concerned early cognition, for which R² increased from 0.218 to 0.365. Most of that increment arose on adding baseline ECs (ΔR² = 0.125); early changes added a further 0.022 after baseline ECs. Complexity-adjusted R² for early cognition was 0.088, 0.189 and 0.167 in M1, M2 and M3, respectively. Thus, the extra early-change terms increased raw in-sample fit without improving adjusted fit beyond the baseline-EC model (Supplementary Table 9).

In M3, more positive baseline IC02 SMG–VAN → IC00 SMN connectivity was associated with greater early cognitive improvement (β = 0.393; pointwise 95% CI, 0.002–0.784; nominal P = 0.049), whereas baseline IC02 SMG–VAN → IC18 MFG–VAN/FPN connectivity showed an inverse association (β = −0.456; −0.849 to −0.062; P = 0.023). Both retained their respective signs in all 43 leave-one-participant-out refits, with coefficient ranges of 0.186–0.588 and −0.541 to −0.383 (Fig. 6). No other baseline EC–clinical association, and none of the eight early-change–clinical associations, had a pointwise 95% interval excluding zero (Fig. 5b; Supplementary Tables 10 and 11).

**Fig. 6.**
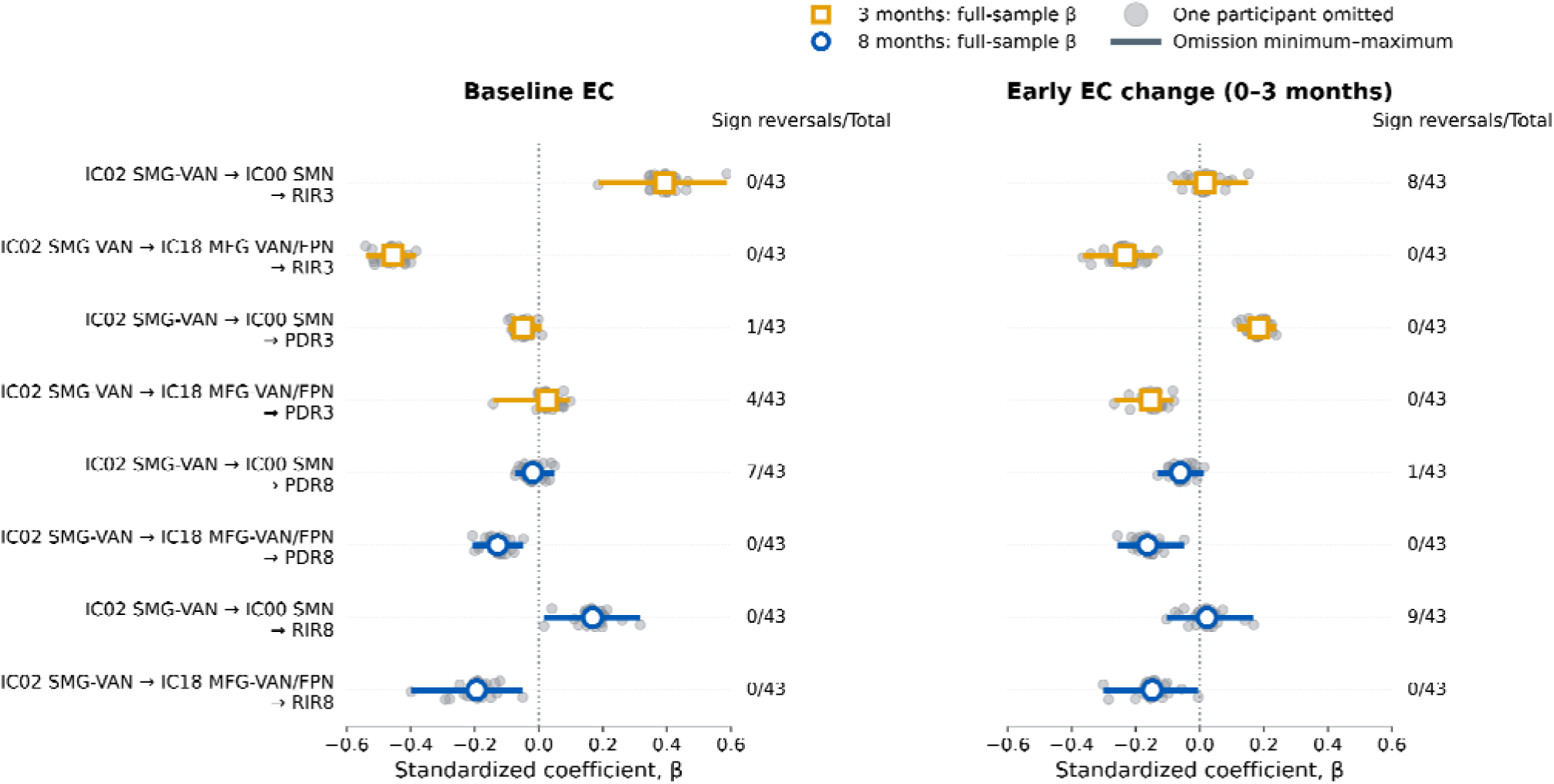
Leave-one-participant-out influence on neural–clinical associations. M3 was refitted after omitting each of 43 participants in turn (22 meditation, 21 control in the full sample; 42 participants per refit). All 43 refits completed. The model specification, two focal connections, covariate adjustment, input standardization and coefficient rescaling were retained at their full-sample definitions. The panels show all 16 neural–clinical associations: baseline EC on the left and early month-3 minus baseline EC change on the right, for IC02 SMG–VAN → IC00 SMN and IC02 SMG–VAN → IC18 MFG–VAN/FPN. Grey points are participant-omission coefficients, vertically offset for visibility; coloured lines span their minima and maxima. Open gold squares and blue circles mark full-sample standardized β for month-3 and month-8 outcomes. Dotted lines mark zero. Counts give opposite-sign refits relative to the full-sample estimate out of 43 omissions. Both baseline-EC associations with early cognition retained direction across all omissions. Omission ranges describe participant influence. PDR, PANSS decrease rate; RIR, RBANS increase rate; EC, effective connectivity.

The full neural model retained a positive treatment coefficient for early IC02 → IC00 change (β = 0.264; 95% CI, 0.040–0.488; P = 0.021); the corresponding IC02 → IC18 estimate was less precise (β = 0.220; −0.011– 0.451; P = 0.062). Treatment-related modulation of connectivity therefore coexisted with limited evidence that these early changes were directly associated with symptom or cognitive benefit in the joint model. An exploratory complementary PEB analysis identified a concurrent early proportional-symptom correspondence for IC02 → IC00 but answered a different question and was limited by clinical-definition and model-family dependence (Supplementary Results and Tables 12–14).

## Discussion

This study links the timing of clinical improvement during meditation-based treatment for schizophrenia to two distinguishable aspects of neural organization: the connections that respond to treatment and the baseline connectivity associated with cognitive improvement. Eight directed connections had shared support across PEB models with and without motion adjustment. The most consistent early effects shared a supramarginal– frontal source and involved its coupling with sensorimotor and bilateral middle-frontal components. Incorporating these connections into the clinical CLPM added the most information about early cognition, principally through their baseline values. These findings identify treatment-responsive network interactions while suggesting that the initial configuration of those interactions provides a complementary context for cognitive response.

The clinical time course motivates this distinction. The symptom treatment contrast increased over follow-up, whereas cognitive benefit was clearest at month 3 and was estimated less precisely at month 8. Early cognitive improvement was positively associated with cumulative later symptom improvement under native CLPM inference; the participant-bootstrap interval and the indirect-effect interval included zero. The resulting hypothesis is that cognitive and symptom responses may be temporally related without a simple sequential mediation pathway adequately capturing that relationship. This extends the parent trial’s efficacy findings by characterizing the timing and interplay of response domains. [11] Meditation research likewise distinguishes relapse prevention, anxiety reduction, distress associated with voices and cognitive performance as different therapeutic outcomes. [1, 2, 10, 32] Their time courses and relevant neural processes may differ.

The imaging findings identify a level of functional organization at which treatment responses were detectable. Intrinsic spatial expression and temporal properties showed no contrasts surviving FDR correction, whereas hierarchical DCM supported changes in directed coupling. The two focal early contrasts remained positive with credible intervals excluding zero across five specifications addressing motion adjustment, temporal representation and visit availability. This consistency strengthens their interpretation as treatment-responsive interactions. It also helps distinguish the contribution of different measurements: a combined analysis of two mindfulness trials found no structural changes despite earlier reports of such effects, illustrating why results from one imaging measure cannot be assumed to generalize to another. [33] Here, the positive connectivity findings motivate an account centred on coordination among brain networks, while the intrinsic-property estimates define the evidence available for changes within components.

The DCM nodes in this study represent distributed functional components. ICA summarizes spatial patterns associated with component time courses, and anatomical labels identify their dominant territories rather than reducing each component to a single cortical locus. [16] IC02 contains supramarginal and frontal territories, with its strongest positive support affiliated predominantly with the ventral attention network. IC00 is predominantly sensorimotor, whereas IC18 contains bilateral middle-frontal territories with both ventral-attention and frontoparietal affiliations. The focal findings therefore concern directed interactions between these distributed systems. This anatomical framing is especially relevant to meditation, which repeatedly engages attention to bodily or sensory experience and the recognition and redirection of distraction. [14, 15, 17, 18] IC02 was the source of three of the eight selected connections, including both consistently supported early effects. This common-source pattern provides a focused hypothesis: supramarginal–frontal coordination with sensorimotor and frontal systems may be particularly responsive to the demands of meditation-based rehabilitation. Task experiments have separated awareness of mind wandering, attentional shifting and sustained focus, while training studies have implicated attentional regulation during emotional processing. [17, 19] Related resting-state studies identify sensory–attention and frontal–parietal changes. [22, 24] These findings make a coordinating role biologically plausible. The present source pattern identifies a candidate component involved in network coordination; a formal hub role would require broader evidence about connectivity centrality or coordination across tasks and networks.

The full eight-connection pattern extends this account beyond the focal pair. It comprises three descriptive patterns: outgoing modulation from IC02 towards sensorimotor, visual and frontal components; target-dependent modulation from the medial visual component towards sensorimotor and precuneus components; and converging inputs to the medial-prefrontal default-mode component from superior-parietal and frontal components. The estimated treatment contrasts have different signs and time courses, suggesting selective reconfiguration rather than a uniform increase in coupling. Several individual visit contrasts remain imprecise even though their temporal terms have strong PEB support. Together, the connections span sensory–motor, attention/control and default-mode systems. This distribution can be considered alongside established sensory-to-transmodal organization of cortical connectivity, while the present analysis specifically characterizes directed inter-component interactions. [34] It motivates testing how sensory, attentional and internally oriented processes are coordinated during treatment.

The temporal profiles offer a further mechanistic clue. Both focal connections were selected for their group-by-quadratic terms, and their estimated treatment contrasts were larger at month 3 than at month 8. Their fitted trajectories describe an estimated early-peaking pattern of coupling reconfiguration. A longitudinal mindfulness trial similarly found an immediate posterior cingulate–prefrontal connectivity effect that was not maintained at later follow-up. [26] One plausible account is that early practice places greater demands on attentional reorientation and sensory–cognitive coordination, followed by more efficient use of those systems as skills develop. Temporary compensation or rebalancing is a more specific version of this hypothesis: it would require showing what functional difficulty is compensated for and how the neural response benefits performance. The present three-visit observations establish temporal modulation, but not yet a homeostatic mechanism. All visits occurred during the intervention period. More frequent assessments of practice, task performance and connectivity could distinguish acquisition-related demands from maintained adaptations.

The neural–clinical models clarify where these connections contribute to understanding clinical response. For month-3 RBANS improvement, baseline EC increased conditional in-sample R² by 0.125, and early EC changes added a further 0.022. The combined increments were smaller for early and later symptom improvement and for later cognition. Complexity-adjusted R² for early cognition was highest after baseline EC entry. More positive baseline IC02→IC00 coupling and less positive baseline IC02→IC18 coupling were associated with greater early cognitive improvement, with coefficient signs retained across all participant omissions. The opposing conditional associations suggest that the initial configuration across connections may be more informative than a general notion of stronger connectivity. These were common associations across treatment arms, identifying neural context for improvement in the trial cohort.

A useful working hypothesis is that initial sensory–attention coordination influences how effectively a person can engage with rehabilitation, while treatment changes some of those same interactions during practice. Contemporary cognitive accounts distinguish increased capacity from improved efficiency through reduced interference and more selective allocation of existing resources. [28] The latter provides a testable interpretation of the baseline–cognition findings. Quantitative reviews also indicate that meditation-related cognitive benefits vary by domain and comparator. [12, 13] RBANS total performance cannot resolve these operations individually. A subsequent trial could relate the two baseline connections to distractor interference, attentional reorienting and sustained focus, and test whether changes in those processes precede broader cognitive improvement.

The correspondence between early EC change and clinical improvement was less clearly resolved in the focused CLPM. This helps locate the next experimental question. Resting connectivity, task operations and changes in clinical scales sample different levels of the treatment response. Studies of rumination and pain illustrate how more proximal outcomes can be linked to neural changes: mindfulness-based cognitive therapy altered connectivity during rumination in association with regulation of attention to bodily sensations, and pain experiments distinguished meditation-related neural responses from placebo and sham conditions. [35–38] Our complementary clinical PEB, which retained DCM uncertainty, identified an early IC02→IC00 correspondence with proportional symptom improvement. Its dependence on clinical representation and model family makes it a focused supplementary observation. The broader result is that treatment-responsive connectivity and baseline neural contributions to cognition are distinguishable, potentially complementary parts of a process model.

The contribution beyond previous functional and structural reports from this trial is the integration of directed interactions among distributed networks with cognitive and symptom trajectories, separating baseline coupling from early connectivity change. [11, 29–31] The workflow connects reproducible component identification, treatment contrasts and clinical correspondence, while documenting what changes across alternative specifications. Its main value is a specific account of which network interactions respond and which features of those interactions add clinical information. The common-source and temporal patterns provide concrete targets for experimental follow-up.

Several features bound these conclusions. This secondary analysis was exploratory, and connection selection and clinical modelling used overlapping participants. Neural–clinical intervals are pointwise and do not incorporate the selection history. The joint models contained 43 participants; repeated scans characterize trajectories but do not add independent patients to between-person associations. Conditional in-sample R² measures description of the observed outcomes, and omission refits characterize participant influence. Analysis-specific measurement availability may affect representativeness; uncertainty in estimated DCM parameters and residual motion-related effects may also affect the associations. The clinical CLPM outcomes are cumulative changes sharing a baseline reference. Prospective confirmation should specify the two connections and clinical contrasts in advance, retain the baseline/change distinction, and assess the proposed attentional processes directly.

The programme was intensive, supervised and embedded in rehabilitation for male inpatients with chronic schizophrenia. Its application to other settings will require attention to practice, engagement, cognitive profile and adverse experiences, which vary across meditation programmes. [7, 39] The immediate biological contribution is the identification of a shared-source network configuration with an estimated early-peaking response, alongside baseline connectivity that adds information about cognitive improvement. This distinction offers a practical design principle for the next trial: measure the neural conditions for engaging with treatment separately from the neural adaptations that accompany it.

## Methods

### Study design, participants and intervention

We performed a secondary analysis of an eight-month, single-centre, parallel-group randomized trial at the First Minzheng Mental Health Center, Shanghai, China (ChiCTR1800014913). [11] Sixty-four male Han Chinese inpatients were assigned in a 1:1 ratio to a meditation-based programme or general rehabilitation. Participants had DSM-IV schizophrenia, chronic illness and persistent positive symptoms, and an MMSE score of at least 24. Allocation used computer-generated random numbers, with enrolment, randomization and allocation performed by different assistants. Outcome assessors were masked to assignment; participants and instructors were not. The ethics review committee of the First Minzheng Mental Health Center approved the study (YJZXLL2017005), and participants provided written informed consent. Detailed trial procedures and eligibility criteria are described in the parent report and Supplementary Methods [11].

From Monday to Saturday, the meditation arm received 30 min of clinician-guided practice and 60 min of general rehabilitation, and the control arm received 90 min of general rehabilitation. Practice included posture, abdominal breathing, attention to the breath and non-reactive awareness of thoughts. Rehabilitation included activities such as manual work, gardening, painting and reading. Both arms also received group sessions with arm-specific content, initially weekly and subsequently monthly. Antipsychotic treatment continued unchanged throughout the whole trial under clinical supervision. Antipsychotic dose was expressed in chlorpromazine equivalents. Clinical assessments and MRI were scheduled at baseline and after 3 and 8 months during the treatment period.

Each analysis included all participants with the measurements required for its model. The clinical analyses used 62 participants with complete required clinical inputs across baseline, month 3 and month 8, without imputation. Three-visit treatment PEB included the 44 participants with fitted DCMs at all three visits; the joint neural–clinical CLPMs included 43 of these participants with complete clinical and motion inputs. One participant in the three-visit PEB sample lacked baseline RBANS. All four neural–clinical CLPMs used these same 43 participants. Analysis-specific eligibility and arm counts are given in Supplementary Table 1.

### Clinical outcomes and average treatment effects

Symptoms were assessed using the Positive and Negative Syndrome Scale (PANSS), and cognition using the Repeatable Battery for the Assessment of Neuropsychological Status (RBANS). [40, 41] Baseline-to-visit improvement was defined as PANSS decrease rate (PDR) = (PANSS − PANSS )/(PANSS − 30), and RBANS increase rate (RIR) = (RBANS − RBANS )/RBANS . Both equal zero at baseline by construction; positive values indicate improvement. Percentage displays multiply these ratios by 100. Mindfulness improvement, assessed with the Five Facet Mindfulness Questionnaire, was summarized in the Supplementary Information.

The time-specific causal estimand was the average difference in improvement under assignment to the two programmes, E[R (1) − R (0)]. Linear mixed-effects models included treatment group, categorical visit, their interaction and the corresponding baseline raw score, with a participant-specific random slope for elapsed months and no random intercept. Models were fitted by restricted maximum likelihood using the Powell optimizer. ATEs were estimated as the treatment-by-visit contrasts relative to the fitted baseline arm difference, with model-based 95% CIs and two-sided Wald tests. Treatment contrasts are interpreted in percentage points. Figure annotations retain percentage notation for these absolute contrasts multiplied by 100. Causal interpretation relies on random assignment and on the assumptions required for the complete-case analysis to represent the target population. Estimated marginal means were evaluated at the pooled mean baseline score. Follow-up-only, participant-clustered and additional-baseline-adjustment analyses are detailed in the Supplementary Methods.

### Clinical cross-lagged panel model and exploratory mediation

A cross-lagged panel model (CLPM), fitted within a structural equation modelling framework, represented PDR and RIR at months 3 and 8. Each month-8 outcome depended on both month-3 outcomes. Treatment, baseline PANSS, baseline RBANS and baseline chlorpromazine-equivalent dose predicted all four outcomes. Within-visit residual covariances were estimated between symptom and cognitive outcomes. All eight input variables, including the treatment indicator, were standardized before estimation. The model was fitted with semopy 2.3.11 using its maximum-likelihood Wishart objective, SLSQP optimization and native expected-information standard errors. Path tests were nominal two-sided Wald tests without multiplicity adjustment.

Exploratory mediation analysis estimated the product of the treatment-to-RIR3 coefficient, conditional on baseline covariates, and the RIR3-to-PDR8 coefficient, additionally conditional on treatment and PDR3. Corresponding regression equations were jointly resampled at the participant level 10,000 times. A 95% percentile interval summarized the standardized indirect-effect estimate, holding the original scaling fixed.

### Imaging acquisition and consensus-guided longitudinal network workflow

MRI was acquired on a Siemens 3-T Verio scanner with a 32-channel head coil. Resting-state echo-planar images had a repetition time of 1.5 s, echo time of 30 ms, 3-mm isotropic resolution and 240 volumes. T1-weighted MPRAGE images had 1-mm isotropic resolution, repetition time 2,530 ms and echo time 3.65 ms. Images were processed with fMRIPrep 22.1.1. [42]

Standard-space BOLD images were further processed in Nilearn 0.12.0. After removal of the initial 10 volumes, nuisance regression included 24 motion regressors, five anatomical CompCor components and cosine drift terms. Parallel pipelines included or omitted global signal regression. Cleaning applied 6-mm spatial smoothing and signal standardization. Scan eligibility required at least 200 post-dummy volumes without concurrent framewise displacement greater than 0.5 mm and standardized DVARS greater than 1.5. Individual network measures and time courses were extracted from the first-stage cleaned images without global signal regression. Detailed processing and quality-control procedures are provided in the Supplementary Methods.

The consensus-guided longitudinal network workflow linked component identification across preprocessing variants, treatment-EC consistency across specifications and subsequent clinical correspondence. Separate canonical ICA decompositions estimated 20 components per preprocessing variant. [43] Nine components were retained through absolute spatial correspondence of at least 0.60 across variants and visual quality control. Anatomical labels combined localization and overlap with established functional and anatomical atlases. [44, 45] Signed spatial loadings describe component orientation. The source-to-target naming convention for DCM edges was maintained throughout. Intrinsic measures comprised spatial expression (MeanZ), sample entropy, normalized permutation entropy and the detrended fluctuation analysis exponent. Baseline-adjusted between-arm contrasts at months 3 and 8 were estimated with mixed-effects models including age and visit-specific mean framewise displacement. Benjamini–Hochberg correction was applied across nine components separately for each metric and visit, giving eight families. The local-measure analysis included 55 participants (30 meditation and 25 control), contributing 99 baseline-linked follow-up observations. Sample preparation and model specification are reported in the Supplementary Methods.

### Spectral DCM and hierarchical treatment PEB

Spectral DCM represented directed neuronal interactions and haemodynamic transformation of the nine component time series through a generative model of their cross-spectral density. [46] Participant–visit models were fully connected, including self-connections, and were fitted using SPM25 with the acquisition-specific TR and TE. Model parameters and their posterior covariance were propagated to PEB. [47] DCM fitting provided 167 participant–visit models from 63 participants (32 meditation and 31 control); treatment trajectory analyses required all three visits and used the 44 complete participants.

Within each participant, the PEB time design contained an intercept, centred linear time at 0, 3 and 8 months, and a centred quadratic term orthogonalized to linear time. Upper-level models estimated treatment-group differences in these temporal parameters. Mean framewise displacement was included through the motion-adjusted specification. Bayesian model reduction and averaging provided posterior inclusion probabilities for treatment-related temporal coefficients. The reporting set comprised the eight connections selected in both the unadjusted and motion-adjusted specifications. The applied inclusion threshold was 0.95; the eight shared connections all exceeded 0.999. Supplementary Methods and Table 8 specify the five model sensitivities and the additional findings confined to the motion-adjusted selection.

Baseline-to-visit treatment contrasts combined the linear and quadratic coefficients using the actual time-design difference and the full posterior covariance. Group coding was converted to the meditation-minus-control difference. Posterior means and 95% Gaussian CrIs are reported in hertz. Inclusion probabilities from reduction/averaging and intervals for full-model temporal contrasts were treated as distinct summaries. Plotted motion-adjusted trajectories were fitted PEB expectations evaluated at mean motion. Two connections with early contrasts consistently supported across the five specifications were used for focused neural–clinical analyses. Selection and subsequent association analyses used overlapping participants.

We assessed treatment-EC consistency across five specifications addressing motion adjustment, temporal representation and visit completeness. Three-visit models used the same 44 participants with no FD adjustment, all-visit FD, or window-specific FD; the latter retained all three neural visits. Paired-visit models used the same 44 participants and then all eligible early (n = 53) or late (n = 46) pairs, with motion adjustment at the included visits. Full-posterior treatment contrasts and 95% credible intervals were compared. The Supplementary Methods specification guide details the designs, mapped priors and distinct purpose of each comparison.

### Neural–clinical cross-lagged panel models and participant influence

Four models were fitted to the same 43 participants using the clinical CLPM estimation convention. M0 reproduced the clinical model. M1 added mean framewise displacement at baseline and month 3 to early equations, and at all three visits to late equations. M2 added baseline IC02 → IC00 and IC02 → IC18 connectivity to every clinical equation. M3 added month-3 minus baseline change in both connections. In M3, each EC change was also modelled as a function of treatment, baseline clinical covariates, both baseline ECs and early motion; the two change residuals were allowed to covary. An auxiliary equation for month-8 motion accommodated its association with preceding model variables without including future motion in early clinical equations. Exact equations are provided in Supplementary Methods.

All inputs, including treatment, were standardized using the common imaging sample. Standardized path coefficients were obtained from the fitted models; pointwise 95% intervals rescaled native expected-information Wald intervals using fitted standard deviations held fixed. Conditional in-sample R² was calculated from fitted structural coefficients and observed predictors. We report R²(M3) − R²(M1), decomposed sequentially into baseline and early-change increments, together with adjusted R² in the supplement.

For participant influence, each model was refitted after omitting each participant in turn, retaining the specification, feature set and full-sample scaling. All 43 refits per model completed. Ranges and sign changes summarize sensitivity to individual participants. The complementary clinical PEB analysis instead treated neural temporal parameters as outcomes of clinical responses while retaining DCM posterior uncertainty. Its focal correspondence, alternative clinical definitions and whole temporal-block comparisons are presented together in the Supplementary Information.

### Statistical reporting and availability

We report effect estimates and uncertainty alongside nominal frequentist P values or Bayesian posterior summaries appropriate to each model. Local-network contrasts used FDR correction; clinical and neural– clinical Wald tests were uncorrected. Analyses used Python, statsmodels, semopy, Nilearn and SPM25. Software versions are provided in the Supplementary Methods.

## Data availability

Aggregate source data for the supported figure panels are available at https://github.com/Vincent-wq/repo_sz_meditation_share. Underlying participant-level data are available from the corresponding authors upon reasonable request.

## Code availability

Analysis source code and an independent notebook for reproducing the supported numerical figure panels are available in the same repository.

## Acknowledgements

The authors would like to thank all the participants, doctors, nurses and students who were involved in the implementation of this RCT. The authors also would like to thank Dr. Pedro Antonio Valdés-Sosa and Dr. María Luisa Bringas-Vega for their feedback and insights that improved the manuscript and visualization. The authors thank Dr. Hu Chuan-Peng for his suggestions during the 2025 COSN hackathon in Shenzhen, China.

## Author contributions

Qing Wang and Donghong Cui conceptualized the research. Qing Wang analysed the data, prepared the visualizations and drafted the manuscript. Lizhao Du contributed to the initial study concept, data collection and curation, and manuscript writing. Qing Wang, Jialing Sheng, Zhong Sun, Yingying Tang and Donghong Cui provided resources and funding support. Qiao Wang contributed to statistical modelling and interpretation of the results. Yuan Shi coordinated and administered the project. Ting Xue contributed to data collection and curation. All authors reviewed the manuscript and provided comments that informed its revision.

